# Glycaemic control and associated factors among patients living with type 2 diabetes in Kinshasa, Democratic Republic of the Congo: a Cross-sectional study

**DOI:** 10.1101/2023.02.03.23285406

**Authors:** Jean-Pierre Fina Lubaki, Olufemi Babatunde Omole, Joel Msafiri Francis

## Abstract

**Objectives:** To assess the prevalence and factors associated with glycaemic control to inform potential interventions to improve glycaemic control in Kinshasa, Democratic Republic of the Congo.

**Design:** This was a cross-sectional study conducted between November 2011–September 2022. We conducted the selection of the participants through a two-stage sampling process. Participants were asked to complete a structured questionnaire and to provide two millilitres of blood for Hb1AC assay. We performed univariate and multivariable logistic regressions to identify factors associated with poor glycaemic control.

**Setting:** A total of 20 randomly selected primary care facilities in Kinshasa, Democratic Republic of the Congo.

**Participants:** The sample included 620 patients living with type 2 diabetes with a median age of 60 (IQR=53.5-69) years.

**Results:** Most of the study participants were female (66.1%), unemployed (67.8%), having income below the poverty line (76.4%), and without health insurance (92.1%). Two-thirds of the participants (420; 67.6%) had poor glycaemic control. Those participants having taken only insulin (AOR=1.64, 95%CI 1.10 to 2.45) and those on a treatment duration ≥7 years (AOR=1.45, 95%CI 1.01 to 2.08) were associated with increased odds of poor glycaemic control, while being overweight (AOR= 0.47, 95%CI 0.26 to 0.85) and those with uncontrolled blood pressure (AOR=0.65, 95% CI 0.48 to 0.90) were protective for poor glycaemic control.

**Conclusions:** This study confirms that poor glycaemic control is common among patients living with type 2 diabetes in Kinshasa, DRC. There is a need for targeted interventions to improve glycaemic control, including metabolic and clinical comorbidity control, lifestyle modifications, and health system factors.

**SIGNIFICANCE OF THE STUDY:** *What is already known on this topic:*

- Glycaemic control is poor in most of the SSA settings, with glycaemic control ranging from 10–60%.
- Factors associated with glycaemic control are context specific; in the Democratic Republic of the Congo, few studies have investigated poor glycaemic control.

*What this study adds:*

- The extent of poor glycaemic control among patients living with type 2 diabetes is determined in Kinshasa, Democratic Republic of the Congo.
- Identification of factors associated with poor glycaemic control in Kinshasa: taking only insulin and having a treatment duration ≥7 years increased the likelihood of poor glycaemic control, while being overweight and having uncontrolled blood pressure were protective.

*How this study might affect research, practice or policy:*

- The study findings will inform potential interventions to improve glycaemic control in Kinshasa, DRC or similar settings elsewhere.

## INTRDODUCTION

Type 2 diabetes is increasing worldwide^1^ – it is expected that the greatest increase in diabetes prevalence will take place in low- and middle-income countries between 2021 to 2045.^2^ On the African Continent, type 2 diabetes is progressing rapidly due to modifiable risk factors, such as obesity and urbanisation.^2^ In sub-Saharan Africa (SSA), diabetes care faces numerous challenges leading to unmet needs and a greater impact on morbidity and mortality.^3 4^ In the Democratic Republic of the Congo (DRC), the prevalence of diabetes is estimated to be 5.8% for adults aged 20 to 79 years,^4^ with a high proportion of persons living with diabetes in urban areas and the western part of the country.^5 6^ The factors associated with diabetes were being male, aged ≥ 40 years, general and abdominal obesity associated with the elderly, family history of diabetes, and hypertension.^6^

Glycaemic control is the cornerstone of diabetes management, as it delays the occurrence of complications, reduces the cost of care and improves patient quality of life. Nevertheless, the control of diabetes remains a worldwide issue, with only about 50% of the patients living with type 2 diabetes controlled.^7^ In SSA, it is estimated that only one-third of patients living with type 2 diabetes reached target glycaemic levels.^8 9^ The accurate knowledge of factors driving glycaemic control in a particular setting is essential to build up an intervention package to improve glycaemic control. Multiple factors drive glycaemic control in SSA, differing across settings.^8^ A recent systematic review of the studies on glycaemic control found: younger and older age, gender, poor socio-economic conditions, place of living, positive family history of diabetes, longer duration of diabetes, treatment modalities and effects, alcohol consumption, smoking presence of comorbidities or complications, and poor management were associated with poor glycaemic control.^9^ Contrarily, high diabetes health literacy, positive perceived family support, adequate coping strategies, dietary adherence, exercise practice, attendance of follow-up appointments, and medication adherence were associated with good glycaemic control.^9^

In the DRC, very few studies have investigated poor glycaemic control among persons/patients living with type 2 diabetes, as high as 86% in the nearby province of Kwilu^10^ and 79.9% in Kinshasa.^11^ Only a few reported factors were found to be associated with glycaemic control among persons living with type 2 diabetes mellitus in the DRC. Sagastume *et al* ^11^ found older patients (> 40 years) had increased odds of achieving glycaemic control than younger patients (< 40 years), while Blum *et al* ^10^ noticed that abdominal obesity and having a body mass index (BMI) > 25 Kg/m2 have been associated with poor glycaemic control. Moreover, we noticed that no study in our setting had previously assessed the link between psychological factors and poor glycaemic control.

This study aimed to assess the extent of glycaemic control and to broaden the knowledge on factors driving poor glycaemic control among patients living with type 2 diabetes in Kinshasa. The results will serve to build an intervention package to improve glycaemic control among patients living with type 2 diabetes in Kinshasa.

## METHODS

This study is a component of the research project on glycaemic control among patients living with type 2 diabetes in Kinshasa, DRC, for which the study protocol was previously published. ^12^

### Ethics

The researchers declared that they complied with the conditions under which this study obtained approval from the ethics committees of the Protestant University of Congo (reference number: CEUPC 0067; Date: 05/02/2021) and Human Research Ethics Committee (Medical) of the University of the Witwatersrand (reference number: M210308; Date: 26/08/2021). The study was conducted according to the ethical guidelines of the Declaration of Helsinki.

Permission was obtained from the Kinshasa Primary HealthCare Network to conduct the study. Informed consent was obtained from each participant. Data collection was done with strict adherence to local COVID-19 regulations.

### Type of study

This was a cross-sectional study among patients recruited from 20 randomly selected health facilities in Kinshasa, DRC.

### Study setting

Our study was multisite within the city of Kinshasa, the DRC’s capital, with an estimated total population of about 15 million inhabitants spread over an area of 9,965 km^2. 13^ The study was conducted in the health facilities belonging to the Catholic Church and the Salvation Army. With a total of 66 health facilities (1 referral hospital and 65 health centres) distributed across 24 health districts, these organisations own most of the facilities that have integrated diabetes care in primary care.

### Study population

The study population consisted of patients living with type 2 diabetes attending health centres offering diabetes care in the Kinshasa Primary Care Network, with about 7326 patients registered in 2020. The inclusion criteria were age ≥18 years and receiving diabetes treatment for at least six months. The exclusion criteria were pregnancy, difficulty communicating due to mental disability, and refusal to provide consent for the study.

### Sample size estimation

The estimated minimum sample size was computed using Epi info version 7.2.2.2. Assuming that the prevalence of poor glycaemic control was 68%,^14^ with a 95% confidence level and a power of 80%, some 59,2% of patients who had a diabetes duration ≤7 years (unexposed) presented with poor glycaemic control, and 74,4% of those who had a diabetes duration >7 years (exposed) presented with poor glycaemic control. The unexposed to exposed ratio is 0.47.^15^ The minimum estimated sample size was 368. Adjusting for a design effect of 1.5, the calculated sample size of 552 was determined. To account for an estimated 10% non-response rate, the minimum required sample size was 614 patients, rounded up to 620 patients.

### Sampling of participants

The selection of the participants was conducted using a two-stage sampling process. The first stage was the selection of healthcare facilities. As the healthcare facilities have an unequal number of patients, the participants were selected by probability proportional to the patient population size. Using this strategy, we randomly selected 20 healthcare facilities out of a possible 48. In the second stage, a minimum of 31 patients were selected from each selected healthcare facility using systematic sampling. This process ensured each patient had the same overall probability of selection or self-weighting. The sampling was done without replacement.

### Data collection

The data collection process lasted from November 2021–September 2022. For each participant, the research team performed physical and anthropometric measurements. These measurements were taken once by trained staff members on the same portable equipment at all the health facilities. The questionnaires were pre-existing standardised questionnaires translated from English into French and Lingala. The questionnaires were tested before data collection and validated for use. At the end of the data collection, a blood sample was taken from the participant. The questionnaire was administered on a tablet or smartphone using REDCap (Research Electronic Data Capture).^16^ The information about the diabetes disease was verified with information in the patient’s medical records, if available.

### Variables of the study

#### Outcome

The main outcome variable was poor glycaemic control which was defined as HbA1c ≥7%,^17^ which was assayed at the laboratory of the School of Medicine at the Protestant University of Congo in Kinshasa. The analysis was completed using an automated Genuis WP 21B with antibody-based immunoassay method of Cypress Diagnostics.^18^

#### Exposures

The possible determinants for glycaemic control sought from the participants were sociodemographic parameters (age, sex, marital status, religion, education level, occupation, ethnic group, income, health insurance, access to food, distance from place of residence to health centre), lifestyle parameters (smoking, risky drinking), clinical parameters (duration of diabetes, height, weight, body mass index, waist circumference, presence of comorbidities, blood pressure, treatment, duration of treatment), and psychological parameters (adherence to treatment, depression, diabetes distress, social support, self-management, knowledge). Supplementary file 1 detailed the exposures, their measurements, operational definitions, reliability, and references.

### Data analysis

All the analyses were performed using survey data analysis with STATA 17 to account for the study design characteristics. We expressed age as median with interquartile range (IQR), as it was not normally distributed. The other variables were analysed as categorical variables and expressed as frequency (n) and percentage (%). Bivariate analysis was performed to compare uncontrolled versus controlled participants in terms of glycosylated haemoglobin using the Chi-square test/Fisher exact test for categorical variables. We further carried out multivariable logistic regression to assess factors associated with glycaemic control. Age, sex, duration of treatment, and food security were included in the regression model a priori. Other variables with a p-value <0.2 in univariate analysis were also included in the model. The p-value of <0.05 was considered statistically significant.

## RESULTS

A total of 620 participants were included in the study out of a total of 627 invited, accounting for a non-response rate of 1.1%. Table 1 summarises the sociodemographic characteristics of the participants.

**Table 1.**
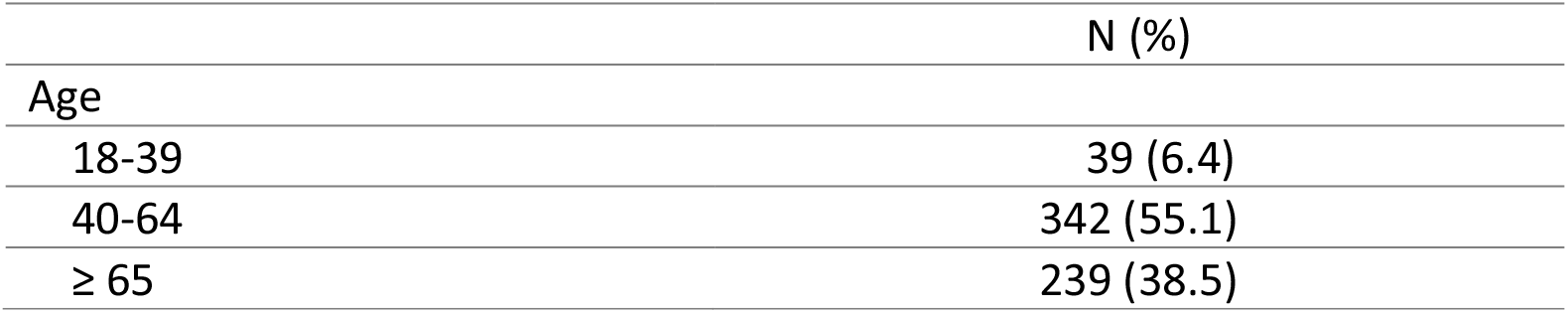
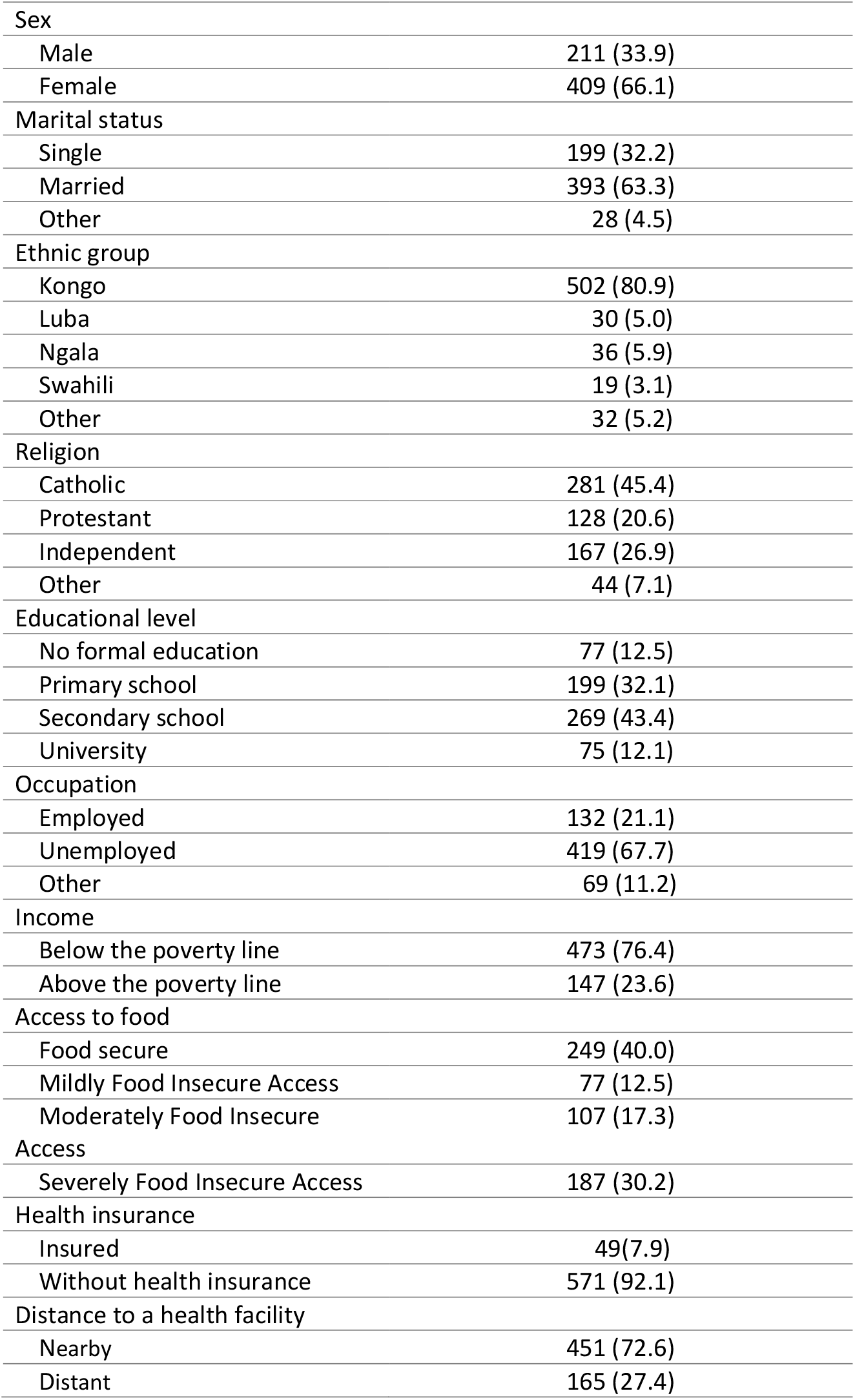
Sociodemographic characteristics of the patients living with type 2 diabetes, Kinshasa, n=620, 2021-2022

### Prevalence of glycaemic control and participants’ characteristics

About two-thirds of the participants (67.8%; n=420) had poor glycaemic control. There was no statistically significant difference between controlled and uncontrolled participants in terms of sociodemographic characteristics (Table 2).

**Table 2.** Sociodemographic characteristics and glycaemic control among patients living with type 2 diabetes in Kinshasa, Democratic Republic of the Congo, n=620, 2021-2022

|  | Controlled<br>(n, %(95%CI)) | Uncontrolled<br>(n, %(95%CI)) | All participants<br>(n, %) | p |
| --- | --- | --- | --- | --- |
| Age |  |  |  | 0.479 |
| 18-39 | 9 (22.8 (10.7 to 42.2)) | 30 (77.2 (57.8 to 89.3)) | 39 (100.0) |  |
| 40-64 | 112 (32.7 (26.7 to 39.4)) | 230 (67.3 (60.6 to 73.3)) | 342 (100.0) |  |
| ≥ 65 | 79 (33.0 (24.8 to 42.2)) | 160 (67.0 (57.6 to 75.2)) | 239 (100.0) |  |
| Sex |  |  |  | 0.989 |
| Male | 68 (32.2 (24.6 to 40.8)) | 143 (67.8 (59.2 to 75.4)) | 211 (100.0) |  |
| Female | 132 (32.2 (25.3 to 40.0)) | 277 (67.8 (60.0 to 74.7)) | 409 (100.0) |  |
| Marital status |  |  |  | 0.413 |
| Single | 68 (34.3 (25.7 to 43.9)) | 131 (65.7 (56.1 to 74.3)) | 199 (100.0) |  |
| Married | 126 (31.3 (25.4 to 39.3)) | 267 (65.7 (60.7 to 74.6)) | 393 (100.0) |  |
| Other | 6 (21.3 (9.7 to 40.6)) | 22 (78.7 (59.4 to 90.3)) | 28 (100.0) |  |
| Ethnic group |  |  |  | 0.405 |
| Kongo | 156 (31.0 (24.9 to 37.9)) | 346 (69.0 (62.1 to 75.1)) | 502 (100.0) |  |
| Luba | 13 (42.0 (25.5 to 60.4)) | 18 (58.0 (39.6 to 74.5)) | 31 (100.0) |  |
| Ngala | 14 (38.7 (26.2 to 52.9)) | 22 (61.3 (47.1 to 73.8)) | 36 (100.0) |  |
| Swahili | 8 (42.0 (24.4 to 61.8)) | 11 (58.0 (38.2 to 75.6)) | 19 (100.0) |  |
| Other | 8 (28.1 (13.5 to 49.5)) | 23 (71.9 (50.5 to 86.5)) | 32 (100.0) |  |
| Religion |  |  |  | 0.201 |
| Catholic | 92 (32.7 (25.7 to 40.7)) | 189 (67.3 (59.3 to 74.3)) | 281 (100.0) |  |
| Protestant | 44 (34.3 (25.4 to 44.6)) | 84 (65.7 (55.4 to 74.6)) | 128 (100.0) |  |
| Independent | 44 (26.2 (16.7 to 38.6)) | 123 (73.8 (61.4 to 83.3)) | 167 (100.0) |  |
| Other | 20 (45.3 (30.5 to 60.9)) | 24 (54.7 (39.1 to 69.5)) | 44 (100.0) |  |
| Educational status |  |  |  | 0.781 |
| No formal | 28 (36.4 (18.2 to 59.7)) | 49 (63.6 (40.3 to 81.8)) | 77 (100.0) |  |
| Primary school | 64 (32.1 (24.6 to 40.6)) | 135 (67.9 (59.4 to 75.4)) | 199 (100.0) |  |
| Secondary school | 87 (32.3 (25.9 to 39.4)) | 182 (67.7 (60.6 to 74.1)) | 269 (100.0) |  |
| University | 21 (27.9 (19.5 to 38.3)) | 54 (72.1 (61.7 to 80.5)) | 75 (100.0) |  |
| Occupation |  |  |  | 0.518 |
| Employed | 45 (34.1 (27.2 to 41.9)) | 87 (65.9 (58.1 to 72.8)) | 132 (100.0) |  |
| Unemployed | 130 (30.9 (24.1 to 38.8)) | 289 (69.1(61.2 to 75.9)) | 419 (100.0) |  |
| Other | 25 (36.3 (25.6 to 48.6)) | 44 (63.7 (51.4 to 74.4)) | 69 (100.0) |  |
| Income |  |  |  | 0.342 |
| Below the poverty line | 149 (31.4 (25.1 to 38.5)) | 324 (68.6 (61.5 to 74.9)) | 473 (100.0) |  |
| Above the poverty line | 51 (34.7 (27.8 to 42.4)) | 96 (65.3 (57.6 to 72.2)) | 147 (100.0) |  |
| Access to food |  |  |  | 0.192 |
| Food secure | 69 (27.6 (19.7 to 37.3)) | 180 (72.4 (62.7 to 80.3)) | 249 (100.0) |  |
| Mildly Food Insecure | 28 (36.5 (24.2 to 50.9)) | 49 (63.5 (49.1 to 75.8)) | 71 (100.0) |  |
| Moderately Food Insecure | 43 (40.2 (31.5 to 49.6)) | 64 (59.8 (50.4 to 68.5)) | 107 (100.0) |  |
| Severely Food Insecure | 60 (31.9 (24.0 to 41.1)) | 127 (68.1 (58.9 to 76.0)) | 187 (100.0) |  |
| Health insurance |  |  |  | 0.782 |
| Insured | 17 (34.5 (20.2 to 52.2)) | 32 (65.5 (47.8 to 79.8)) | 48 (100.0) |  |
| Uninsured | 183 (32.0 (25.8 to 39.0)) | 388 (68.0 (61.0 to 74.2)) | 571 (100.0) |  |
| Distance to a health facility |  |  |  | 0.644 |
| Nearby | 148 (32.8 (25.8 to 40.6)) | 303 (67.2 (59.4 to 74.2)) | 451(100.0) |  |
| Distant | 52 (30.7 (23.7 to 38.7)) | 117(69.3 (61.3 to 76.3)) | 169 (100.0) |  |

However, controlled participants differed significantly from uncontrolled participants in terms of BMI (p=0.005), control of blood pressure (p=0.027), and treatment regimens (p=0.002) (Table 3).

**Table 3.** Lifestyle and clinical characteristics and glycaemic control among patients living with type 2 diabetes in Kinshasa, Democratic Republic of the Congo, n=620, 2021-2022

|  | Controlled<br>(n, %(95%CI)) | Uncontrolled<br>(n, %(95%CI)) | All participants<br>(n, %) | p |
| --- | --- | --- | --- | --- |
| Smoking |  |  |  | 0.268 |
| No | 200 (32.4 (26.5 to 38.9)) | 416 (67.6 (61.1 to 73.5)) | 616 (100.0) |  |
| Yes | 0 (0.0 (0.0)) | 4 (100.0 (0.0)) | 4 (100.0) |  |
| Alcohol disorder |  |  |  | 0.566 |
| No disorder | 198 (32.4 (26.5 to 38.8)) | 413 (67.6 (61.2 to 73.5)) | 611 (100.0) |  |
| Health risk consumption | 2 (22.1 (4.0 to 66.2)) | 7 (77.9 (33.8 to 96.0)) | 9 (100.0) |  |
| Duration of diabetes |  |  |  | 0.549 |
| 0-5 years | 84 (31.3 (23.0 to 41.0)) | 183 (68.7 (59.0 to 77.0)) | 267 (100.0) |  |
| 5-10 years | 76 (35.1 (26.5 to 44.8)) | 141 (64.9 (55.2 to 73.5)) | 217 (100.0) |  |
| ≥10 years | 40 (29.3 (22.2 to 37.5)) | 96 (70.7 (62.5 to 77.8)) | 136 (100.0) |  |
| Body mass index |  |  |  | 0.005* |
| Underweight | 14 (21.6 (13.4 to 33.0)) | 51 (78.4 (67.0 to 86.6)) | 65 (100.0) |  |
| Normal | 102 (29.3 (21.9 to 37.9)) | 245 (70.7 (62.1 to 78.1)) | 347 (100.0) |  |
| Overweight | 66 (46.6 (39.1 to 54.2)) | 76 (53.4 (45.8 to 60.9)) | 142 (100.0) |  |
| Obesity | 18 (27.3 (15.7 to 43.1)) | 48 (72.7 (56.9 to 84.3)) | 66 (100.0) |  |
| Waist circumference |  |  |  | 0.083 |
| Abnormal | 93 (38.2 (29.8 to 47.3)) | 150 (61.8 (52.7 to 70.2)) | 243 (100.0) |  |
| Normal | 107 (28.3 (21.5 to 36.3)) | 270 (71.7 (63.7 to 78.5)) | 377 (100.0) |  |
| Presence of comorbidities |  |  |  | 0.266 |
| Yes | 136 (33.9 (27.6 to 40.7)) | 265 (66.1 (59.3 to 72.4)) | 401 (100.0) |  |
| No | 64 (29.2 (21.6 to 38.1)) | 155 (70.8 (61.9 to 78.4)) | 218 (100.0) |  |
| Blood pressure |  |  |  | 0.027* |
| Controlled | 117 (29.2 (22.9 to 36.4)) | 283 (70.8 (63.6 to 77.1)) | 400 (100.0) |  |
| Uncontrolled | 83 (37.7 (30.8 to 45.2)) | 137 (62.3 (54.8 to 69.2)) | 220 (100.0) |  |
| Treatment |  |  |  | 0.002* |
| Insulin | 87 (26.0 (20.7 to 32.1)) | 247 (74.0 (67.9 to 79.3)) | 334 (100.0) |  |
| Oral hypoglycaemic drugs | 98 (41.3 (31.6 to 51.7)) | 139 (58.7 (48.3 to 68.4)) | 237 (100.0) |  |
| Mixed | 15 (30.7 (21.1 to 42.3)) | 34 (69.3 (57.7 to 78.9)) | 49 (100.0) |  |
| Duration of treatment |  |  |  | 0.229 |
| < 7 years | 132 (34.1 (27.3 to 41.6)) | 255 (65.9 (58.4 to 72.7)) | 387 (100.0) |  |
| ≥ 7 years | 68 (29.1 (21.9 to 37.4)) | 165 (70.9 (62.6 to 78.1)) | 233 (100.0) |  |

\*p<0.05

Perceived support from significant others (p=0.005), perceived family support (p=0.020), diabetes treatment regimen distress (p=0.029), and adherence to physical activity (p=0.017) were statistically significant between controlled and uncontrolled participants (Supplementary table 1).

### Determinants of glycaemic control

Being on insulin (AOR=1.64, 95%CI 1.10 to 2.45) and having a treatment duration ≥7 years (AOR=1.45, 95%CI 1.01 to 2.08) were associated with increased odds of poor glycaemic control. Being overweight (AOR= 0.47, 95%CI 0.26 to 0.85) and having uncontrolled blood pressure (AOR=0.65, 95% CI 0.48 to 0.91) were associated with decreased odds of poor glycaemic control (Table 4).

**Table 4.** Results of survey logistic regression estimating the association between different exposures and poor glycaemic control in 620 patients with type 2 diabetes in Kinshasa, Democratic Republic of the Congo, 2021-2022

|  | Univariate analysis |  | Multivariate analysis |  |
| --- | --- | --- | --- | --- |
|  | p | COR (95%CI) | p | AOR (95%CI) |
| Age (years) |  |  |  |  |
| 18-39 |  | 1 |  | 1 |
| 40-64 | 0.276 | 0.61 (0.24 to 1.54) | 0.652 | 0.81 (0.32 to 2.08) |
| ≥ 65 | 0.271 | 0.60 (0.23 to 1.54) | 0.736 | 0.86 (0.35 to 2.13) |
| Sex |  |  |  |  |
| Male |  | 1 |  | 1 |
| Female | 0.989 | 1.00 (0.64 to 1.55) | 0.494 | 1.16 (0.75 to 1.79) |
| Access to food |  |  |  |  |
| Food secure |  | 1 |  | 1 |
| Mildly Food Insecure | 0.199 | 0.66 (0.35 to 1.27) | 0.079 | 0.59 (0.33 to 1.07) |
| Moderately Food Insecure | 0.008 | 0.57 (0.38 to 0.85) | 0.083 | 0.69 (0.46 to 1.05) |
| Severely Food Insecure | 0.468 | 0.81 (0.45 to 1.46) | 0.670 | 0.90 (0.53 to 1.51) |
| Body mass index |  |  |  |  |
| Underweight |  | 1 |  | 1 |
| Normal | 0.178 | 0.66 (0.36 to 1.22) | 0.288 | 0.73 (0.41 to 1.32) |
| Overweight | 0.000 | 0.32 (0.18 to 0.55) | 0.015* | 0.47 (0.26 to 0.85) |
| Obese | 0.515 | 0.73 (0.28 to 1.94) | 0.574 | 1.21 (0.60 to 2.48) |
| Waist circumference |  |  |  |  |
| Normal |  | 1 |  | 1 |
| Abnormal | 0.084 | 0.64 (0.38 to 1.07) | 0.328 | 0.80 (0.51 to 1.26) |
| Blood pressure |  |  |  |  |
| Controlled |  | 1 |  |  |
| Uncontrolled | 0.027 | 0.68 (0.84 to 0.95) | 0.011* | 0.65 (0.48 to 0.90) |
| Duration of treatment (years) |  |  |  |  |
| < 7 |  | 1 |  | 1 |
| ≥ 7 | 0.229 | 1.26 (0.85 to 1.87) | 0.043* | 1.45 (1.01 to 2.08) |
| Treatment regimen |  |  |  |  |
| OAD |  | 1 |  | 1 |
| Insulin | 0.003 | 2.00 (1.31 to 3.05) | 0.019* | 1.10 (1.10 to 2.45) |
| Mixed | 0.144 | 1.59 (0.84 to 2.99) | 0.319 | 1.39 (0.71 to 2.74) |
| Significant other support |  |  |  |  |
| Low support |  | 1 |  | 1 |
| Moderate support | 0.465 | 0.85 (0.54 to 1.34) | 0.244 | 0.78 (0.51 to 1.20) |
| High support | 0.002 | 0.42 (0.25 to 0.71) | 0.128 | 0.64 (0.36 to 1.15) |
| Family support |  |  |  |  |
| Low support |  | 1 |  | 1 |
| Moderate support | 0.586 | 1.14 (0.69 to 1.91) | 0.533 | 1.22 (0.63 to 2.34) |
| High support | 0.015 | 0.50 (0.29 to 0.86) | 0.359 | 0.70 (0.31 to 1.55) |
| Diabetes distress |  |  |  |  |
| No distress |  | 1 |  | 1 |
| Moderate distress | 0.017 | 1.75 (1.12 to 2.73) | 0.179 | 1.42 (0.84 to 2.39) |
| High distress | 0.009 | 1.72 (0.83 to 3.54) | 0.901 | 0.95 (0.42 to 2.23) |
| Physical activity |  |  |  |  |
| Poor adherence |  | 1 |  | 1 |
| Good adherence | 0.019 | 1.55 (1.08 to 2.22) | 0.067 | 1.34 (0.98 to 1.80) |
| Knowledge on diabetes |  |  |  |  |
| Low |  | 1 |  | 1 |
| Acceptable | 0.004 | 1.45 (1.14 to 1.83) | 0.054 | 0.69 (0.48 to 1.01) |
| Good | 0.846 | 1.04 (0.68 to 1.59) | 0.419 | 1.13 (0.83 to 1.52) |

\*p<0.05

## DISCUSSION

This study was designed to assess the extent of poor glycaemic control among patients living with type 2 diabetes in Kinshasa, including its driving factors. The results found a prevalence of poor glycaemic control of 67.8%. No sociodemographic and lifestyle characteristics were associated with glycaemic control. Taking only insulin and having a treatment duration ≥7 years increased the odds of poor glycaemic control, while being overweight and having uncontrolled blood pressure were protective.

This study found that more than two-thirds of the participants have poor glycaemic control, thus corroborating the findings of other studies in SSA. ^8 9^ Furthermore, this study found a higher prevalence of poor glycaemic control than that found by Blum *et al* in the nearby rural province of Kwilu in the DRC. Findings from this study are lower than those found in the European or North American studies, ^19 20^ and indicated poor diabetes care in Kinshasa. These results also translated the issues in SSA in general, where diabetes care faces multiple barriers. ^3 21^ Moreover, self-management in SSA is poor and represents a threat to the health of individuals and capacity of the health system.^22^ Meanwhile, diabetes is becoming more predominant, with great economic and health impacts. ^3 21^ In a study in the Kinshasa Health Network in 2015, Kapongo *et al* found that the capacities, knowledge, and practice of type 2 diabetes care were poor and stated that the training of healthcare professionals, equipment of health facilities and development of clinical guidelines could help in improving glycaemic control.^23^ Thus, effective funding, and better preparation for diabetes care is crucial.

Most of our participants (93.7%) were older than 40 years. This proportion aligns with the classic description of type 2 diabetic patients, in which the disease appears in individuals older than 40 years most of the time. Female patients represented approximately two-thirds of the participants. A retrospective analysis of the Kinshasa Health Network database conducted by Sagastume *et al*^11^ also found the same-sex prevalence. This high prevalence of type 2 diabetes affects more women than men, due to the higher metabolic risk in the former.^24^ Furthermore, the health-seeking behaviour of women is greater than men.^25^ A distribution of religion and ethnic groups among the participants corresponded linearly to the distributions of those parameters in the general population. Most of the participants were unemployed, poor and without health insurance, which has been representative of the condition of the general population in Kinshasa. No sociodemographic and lifestyle characteristics were associated with poor glycaemic control. Our finding here has been corroborated by the study of Blum *et al*,^10^ who also found no sociodemographic or lifestyle factors associated with poor glycaemic control in a cross-sectional survey in the nearby province of Bandundu in the DRC. We suppose that the tools used to characterise patients were unsuitable for the particularities of our setting. A recent systematic review on glycaemic control among patients living with type 2 diabetes in SSA found that younger and older age, gender, lower income, absence of health insurance, low level of education, and place of residence were associated with poor glycaemic control.^9^ In a retrospective study in Kinshasa, Sagastume *et al*^11^ found that younger patients needed prioritised attention to reach glycaemic targets. Research with more appropriate and suitable tools will be needed to better define the contribution of sociodemographic and lifestyle characteristics on glycaemic control in our setting. Nevertheless, interventions for better glycaemic control have to prioritise vulnerable groups, such as younger and older age, women and non-insured patients. Implementing universal coverage can increase access to care for the aforementioned groups.^26^

In this study, participants taking only insulin were 1,64 times more likely to have poor glycaemic control than those on oral hypoglycaemic drugs. Studies have shown that only around one-fourth of patients on insulin could achieve glycaemic targets because the participants might be erroneously taking an insufficient daily dose and incorrectly titrating insulin.^27^ The lack of adherence to the prescribed regimen has been also a significant factor in poor glycaemic control and was magnified in our setting, as most of the patients were unemployed and not covered by health insurance. They were unable to adequately follow the prescribed regimen when they lacked food. The psychological resistance to insulin, prevalent in our setting according to the study by Rita *et al*,^28^ could also be another explanation for poor glycaemic control among patients living with type 2 diabetes in our study. In the diabetes attitudes, wishes and needs second study (DAWN), participants reported low confidence in the efficacy of insulin, with 26.9% of participants abstaining from insulin because they thought insulin unfeasible or impracticable to manage their diabetes.^29^ Healthcare providers must ensure that psychological resistance to initiating insulin is adequately addressed, and effectively train the patients to correctly follow their prescriptions. Type 2 diabetes is a lifestyle disease, and all guidelines prescribe that insulin therapy should be accompanied by lifestyle modification and oral hypoglycaemic drugs. The prepotency of insulin use highlighted the lack of appropriate guidelines or poor clinicians’ adherence to evidence-based clinical guidelines. Clear management guidelines must also be adapted for the use of available medicines and efforts must be made to offer new hypoglycaemic agents at affordable prices.

In this study, the odds of overweight participants having poor glycaemic control were reduced by 53.0%. Blum *et al*^10^, in their study near Kinshasa, also found that BMI>25 Kg and abdominal obesity were protective against poor glycaemic control. The authors stated that this finding could reflect the existence of special features of diabetes in SSA. Plečko *et al*,^30^ in an analysis encompassing four large international clinical databases for critically ill patients, found that higher BMI in patients with diabetes was associated with lower average glucose. The authors suggested that their finding could be explained by the process of care, as the patients with higher BMIs had more glucose measurements and were receiving higher insulin doses. Our finding contrasted the well-known relationship between being overweight and suboptimal glycaemic control and poor glycaemic control.^31^ Weight loss or the prevention of weight gain is an important goal in the management of type 2 diabetes or prediabetes.^32^ However, increasing weight could also arise in diabetic patients due to the effect of antidiabetic medication on body weight. Apart from metformin and thiazolidinediones, other antidiabetic agents could lead to weight gain.^33^ In Kinshasa, insulin is largely used and there has been a limited range of affordable medications for patients. Healthcare providers must furthermore ensure that the patients are adequately managed to avoid adverse effects.^33^

In the study sample, patients with uncontrolled blood pressure reduced the odds of having poor glycaemic control by 35.0%. Mobula *et al*,^34^ in a Ghanaian study, also found that systolic blood pressure was significantly higher among patients with adequate glycaemic control compared to the group with poor glycaemic control. As discussed by Mobula *et al*,^34^ among patients with good glycaemic control, it can be that there was a significantly higher proportion of patients with dual diagnosis—hypertension and diabetes. This observation can also be explained by the fact that health providers give more attention to persons with comorbidity or an increase in healthcare utilisation by the persons with comorbidity.^35^ Hypertension is frequently associated with diabetes,^36^ which indicates that more insight into adequate management of hypertension among patients with diabetes in our setting will be required.^35^

This study found that a treatment duration ≥7 years increased the odds of poor glycaemic control by 1,45 times. Longer duration of treatment has been linked to poor glycaemic control in SSA.^37^ As diabetes takes longer to emerge, there is a progressive deterioration of the pancreas function, therefore requiring more adjustments in the treatment of older patients, who can develop comorbidities.^38^ One may note that in SSA, there has been a high proportion of undiagnosed diabetes, and usually at the time of diagnosis made the disease is relatively advanced with complications already present.^39^ Health providers must be informed of the progression of diabetes and be able to adjust the treatments for patients accordingly.

### Limitations of the study

This study estimated the extent of poor glycaemic control among type 2 diabetes patients in Kinshasa. Also, we have broadly assessed psychological scales regarding glycaemic control in SSA to promote understanding in the management of patients with diabetes. Because of the cross-sectional nature of the study, it is not possible to ascertain a causal relationship between poor glycaemic control and the determinants. Other potential biases include selection (non-response of the eligible participants), recall, interviewer, and social desirability. These biases were minimised by ensuring effective training of the data collectors to make certain that the aim and objectives of the study were clearly stated to the participants, and that questions asked were communicated with a non-judgemental attitude.

## CONCLUSION

This study confirms that poor glycaemic control is common among patients living with type 2 diabetes in Kinshasa, DRC. There is a need for targeted interventions to improve glycaemic control, including metabolic and clinical comorbidity control, lifestyle modifications, and health system factors.

## Supporting information

Exposures' measurements tools

Glycaemic control and psychological characteristics

## Data Availability

All datasets supporting the conclusions in this article are included within the article

## Special acknowledgement

The authors would like to thank all the staff of the Kinshasa Primary Health Network for their support during the study. We thank Mrs Manase Lusuami for performing the laboratory assays, and Christian Mungongo Kifu for his support during data collection and data management.

## Author Contributions

JPF (guarantor), OBO, and JMF designed the study. JPF contributed to acquisition of funding and data. JPF oversaw the research process. JPF and JMF performed the data processing and quality control. JPF conducted the statistical analyses, drafted the manuscript, and is the guarantor of this work. JPF, OBO, and JMF interpreted the data. All authors edited and critically reviewed the manuscript for intellectual content and approved the final version.

## Funding support

The authors received no specific funding for this work. The Protestant University of Congo provided laboratory reagents and facilitated laboratory assays.

## Declaration of interests

The authors declare that they have no competing interests.

## Patient consent for publication

Informed consent was obtained from each participant.

## Data availability statement

All datasets supporting the conclusions in this article are included within the article.

## Disclaimers

The authors stated that the views expressed in this article are their own and not an official position of the institution or funder.

## Notes

### Competing Interest Statement

The authors have declared no competing interest.

### Author Declarations

Ethics committees of the Protestant University of Congo (reference number: CEUPC 0067; Date: 05/02/2021) and Human Research Ethics Committee (Medical) of the University of the Witwatersrand (reference number: M210308; Date: 26/08/2021).

