## Supplementary material for "Glycaemic control and associated factors among patients living with type 2 diabetes in Kinshasa, Democratic Republic of the Congo: a Cross-sectional study": Exposures' measurements tools

**Supplementary file 1 Measurement tools, operational definitions, and references for the exposures**

**Sociodemographic parameters**

We considered a distance from the participant's place of residence to the health centre of < five kilometres or < one hour as nearby, while > five kilometres or > one hour as distant. A daily income < 3800 Congolese Francs (< US $1.9) was set as the threshold for poverty.[1] We measured food security status with the Household Food Insecurity Scale (HFIAS), a nine-item measurement tool. It reported the experience of the past four weeks.[2] The HFIAS showed a good internal consistency (Cronbach's alpha of 0.76 and 0.73 in two rounds) in an Ethiopian study and the authors concluded that the tool was simple but an adaptation of questions and wording, as well as additional examples before the application could be necessary.[3] Our sample data displayed a good internal consistency (Cronbach's alpha: 0.96). The HFIAS categorises households into four levels of food access: food secure, mild food insecure, moderately food insecure, and severely food insecure.[4]

**Clinical parameters**

For physical measurements, trained staff members measured clinical parameters (height, weight, waist circumference, and blood pressure) using validated equipment. The height of the participant was measured with a stadiometer on which the participant stands bare feet with his or her back against the vertical scale, feet parallel to each other, toes pointing forward and the soles of feet flat on the floor. The measurement was read on the vertical scale to the nearest centimetre. The weight was assessed using a SECA scale 260 with the participant wearing light clothing, in bare feet, minus outer clothing, no heavy jewellery, and empty pockets. The weight was expressed in kilograms. The waist circumference was measured with the patient standing upright with feet slightly apart and arms hanging loosely on each side. A tape was placed just above the hip bones to measure the middle and was taken just after the participant breathed out, to the nearest centimetre.[5]

The BMI was calculated as weight(Kg)/height(squared meters) and was categorised as follows: underweight (BMI<18.5 Kg/$m^2$), normal weighted (18.5-24.99 Kg/$m^2$), overweight (BMI≥25 Kg/m2), and obese ≥30Kg/m2.[6] For the waist circumference, normal values were defined according to sex: men <102 centimetres; and women <88 centimetres.[7] Controlled blood pressure was defined as a systolic blood pressure level of 140 mmHg or higher or lower and/or a diastolic blood pressure level of 90 mmHg or lower.

We have divided the duration of diabetes into three categories: less than 5 years, 5 to 10 years, and 10 years or more. The duration of the treatment has two categories: less than 7 years, and 7 years or more. We considered three options for treatment regimens: insulin alone, oral hypoglycaemic drugs, and mixed treatment (insulin plus oral hypoglycaemic drugs).

**Psychological parameters**

We assessed knowledge of diabetes with the Simplified Diabetes Knowledge Scale, a true/false response format of the Revised Diabetes Knowledge Scale of the Michigan Diabetes Research Center.[8] 9 It contained 20 items with three related to insulin users. It has an internal reliability of 0.71, an item correlation of 0.26–0.58, and corrected item-total correlation of >0.2 for all items, except for items 7, 8, and 20. [8] Our sample data demonstrated a similar Cronbach's alpha at 0.74.

The Simplified Diabetes Knowledge Scale was used as a continuous score. For analysis, the score was categorised into three categories, namely low (<7), acceptable (7–10), and good (≥11).

We assessed treatment adherence by using the four-item Morisky Green Levine (MGL) test.[10] The four-item MGL test has an alpha reliability of 0.61. Our data showed similar Cronbach's alpha of 0.64. The scale is a yes/no scale with 'yes' coded as 1, and 'no' as 0. The MGL test divides patients into adherence categories: high, medium, and low with 0, 1–2, and 3–4 points, respectively.

We assessed social support by applying the Multidimensional Scale for Perceived Social Support (MSPSS).[11] The MSPSS is a brief self-report questionnaire that contains 12 items rated on a five-point Likert-type scale. The MSPSS assessed three subscales of social support: family, friends, and significant others. The internal consistency of the total scale was 0.87: the subscales were 0.84 (friends), 0.85 (family), and 0.74 (significant others).[11] Our sample data showed a similar Cronbach's alpha of 0.89. For the MSPSS, the mean scores were calculated as follows: Significant Others Subscale - sum across items 1, 2, 5 & 10, divided by 4; Family Subscale - sum across items 3, 4, 8 & 11, divided by 4; and Friends Subscale - sum across items 6, 7, 9 & 12, divided by 4. The total scale was calculated as the sum across all 12 items, divided by 12. Any mean scale score ranging from 1–2.9 could be considered low support, a score of 3–5 as moderate support, and a score from 5.1–7 as high support.

We conducted a screening for depression with the Patient Health Questionnaire-9 (PHQ-9).[12] The PHQ-9 is the nine-point depression module of the full PHQ and is a screening tool and measure of depression severity. The Cronbach's alpha was 0.89 in a primary healthcare study.[13] Our sample demonstrated a Cronbach's alpha of 0.77, less than that of the previous study but acceptable. Depression screening with PHQ-9 determined the following categories: moderate depression (12–14), moderately severe depression (15–19), and severe depression (20–27).[12]

We measured diabetes distress using the Diabetes Distress Scale (DDS),[14] a 17-item scale that captures four critical dimensions of distress: emotional burden (five items), regimen distress (five items), interpersonal distress (three items), and physician distress (four items). The internal consistency expressed by Cronbach's α was 0.93 for the total score, 0.88 for emotional burden, 0.88 for physician-related distress, 0.88 for regimen-related distress, and 0.88 for interpersonal-related distress scales.[14] Our sample showed a Cronbach's alpha of 0.94. For the DDS and its subscales, a mean item score of 2.0–2.9 are considered 'moderate distress,' while a mean item score >3.0 is considered 'high distress'.

We determined diabetes self-management with the use of the Diabetes Self-Management Questionnaire (DSMQ), a 16-item questionnaire comprising four subscales: dietary control (four items), glucose management (five items), physical activity (three items), and physician contact (three items). Low adherence was defined as a score of dietary control <4, a score of glucose management <5, a score of physical activity <3, and a score of physician contact <3.[15] The overall internal consistency (Cronbach's alpha) was good (0.84) and consistencies of the subscales were acceptable (glucose management [0.77]; dietary control [0.77]; physical activity [0.76]; and physician contact [0.60]).[15] Our sample showed an acceptable Cronbach's alpha of 0.76.

**Lifestyle parameters**

For lifestyle parameters, we investigated substance use. We tested alcohol use with the Alcohol Use Disorders Identification Test-Concise (AUDIT-C), where a score ≥4 (in men) or ≥3 (in women) represents health-risk consumption.[16] The internal consistency of AUDIT-C was 0.75, and its test-retest reliability was 0.93.[17] Our sample demonstrated a Cronbach's alpha of 0.64. The history of smoking was sought out from the participants and focused only on current smoking.
