## Supplementary material for "Glycaemic control and associated factors among patients living with type 2 diabetes in Kinshasa, Democratic Republic of the Congo: a Cross-sectional study": Glycaemic control and psychological characteristics

**Supplementary table 1** Psychological characteristics and glycaemic control among patients living with type 2 diabetes in Kinshasa, Democratic Republic of the Congo, n=620 (2021-2022)

| | **Controlled (n, %(95%CI))** | **Uncontrolled (n, %(95%CI))** | **All participants (n, %)** | **p** |
|---|---|---|---|---|
| Adherence to treatment | | | | 0.427 |
| High | 110 (31.3 (23.4 to 40.3)) | 241 (68.7 (59.7 to 76.6)) | 351 (100.0) | |
| Moderate | 76 (35.0 (29.2 to 41.4)) | 141 (65.0 (58.6 to 70.8)) | 217 (100.0) | |
| Low | 14 (26.8 (17.0 to 39.7)) | 38 (73.2 (60.3 to 83.0)) | 52 (100.0) | |
| Depression | | | | 0.535 |
| Without depression | 198 (32.3 (26.5 to 38.7)) | 414 (67.6 (61.3 to 73.5)) | 612 (100.0) | |
| Moderate depression | 1 (16.7 (2.6 to 60.0)) | 5 (83.3 (40.0 to 95.4)) | 6 (100.0) | |
| Moderately severe depression | 1 (50.1 (4.6 to 95.4)) | 1 (49.9 (4.6 to 95.4)) | 2 (100.0) | |
| Multidimensional perceived social support | | | | |
| Total score | | | | 0.299 |
| Low support | 40 (28.8 (20.6 to 38.6)) | 99 (71.2 (61.4 to 79.4)) | 139 (100.0) | |
| Moderate support | 150 (32.5 (25.6 to 40.4)) | 310 (67.5 (59.6 to 74.4)) | 460 (100.0) | |
| High support | 10 (47.4 (27.8 to 67.8)) | 11 (52.6 (32.2 to 72.2)) | 21 (100.0) | |
| Significant others | | | | 0.006* |
| Low support | 49 (26.9 (19.3 to 36.1)) | 133 (73.1 (63.9 to 80.7)) | 182 (100.0) | |
| Moderate support | 98 (30.2 (23.2 to 38.2)) | 227 (69.8 (61.8 to 76.8)) | 325 (100.0) | |
| High support | 53 (46.7 (38.1 to 55.6)) | 60 (53.3 (44.4 to 61.9)) | 113 (100.0) | |
| Family | | | | 0.020* |
| Low support | 44 (31.5 (24.7 to 39.3)) | 96 (68.5 (60.7 to 75.3)) | 140 (100.0) | |
| Moderate support | 111 (28.7 (21.4 to 37.2)) | 275 (71.3 (62.8 to 78.6)) | 386 (100.0) | |
| High support | 45 (47.8 (36.3 to 59.5)) | 49 (52.2 (40.5 to 63.7)) | 94 (100.0) | |
| Friends | | | | 0.463 |
| Low support | 100 (35.3 (27.1 to 44.5)) | 182 (64.7 (55.5 to 72.9)) | 282 (100.0) | |

| | | | | |
|---|---|---|---|---|
| Moderate support | 95 (29.6 (20.7 to 40.4)) | 226 (70.4 (59.6 to 79.3)) | 321 (100.0) | |
| High support | 5 (29.4 (17.1 to 45.6)) | 12 (70.6 (54.4 to 82.9)) | 17 (100.0) | |
| Diabetes distress | | | | |
| Total score | | | | 0.113 |
| No distress | 105 (39.0 (31.8 to 46.8)) | 163 (61.0 (53.2 to 68.2)) | 268 (100.0) | |
| Moderate distress | 37 (26.8 (19.5 to 35.5)) | 101 (73.2 (64.5 to 80.5)) | 138 (100.0) | |
| High distress | 58 (27.2 (16.9 to 40.6)) | 156 (72.8 (59.4 to 83.1)) | 214 (100.0) | |
| Emotional | | | | 0.375 |
| No distress | 135 (34.1 (27.9 to 40.9)) | 260 (65.9 (59.1 to 72.1)) | 395 (100.0) | |
| High distress | 65 (28.9 (19.8 to 40.2)) | 160 (71.1 (59.8 to 80.2)) | 225 (100.0) | |
| Regimen | | | | 0.033* |
| No distress | 102 (39.7 (32.1 to 47.9)) | 154 (60.3 (52.1 to 67.9)) | 256 (100.0) | |
| Moderate distress | 31 (21.5 (14.7 to 30.3)) | 113 (78.5 (69.7 to 85.3)) | 144 (100.0) | |
| High distress | 67 (30.5 (20.7 to 42.5)) | 153 (69.5 (57.5 to 79.3)) | 220 (100.0) | |
| Interpersonal | | | | 0.293 |
| No distress | 91 (36.7 (29.1 to 45.1)) | 156 (63.3 (54.9 to 70.9)) | 247 (100.0) | |
| Moderate distress | 37 (28.4 (22.3 to 35.4)) | 93 (71.6 (64.6 to 77.7)) | 130 (100.0) | |
| High distress | 72 (29.6 (19.9 to 41.6)) | 169 (70.4 (58.4 to 80.1)) | 243 (100.0) | |
| Physician | | | | 0.134 |
| No distress | 106 (38.0 (30.8 to 45.8)) | 172 (62.0 (54.2 to 69.2)) | 278 (100.0) | |
| Moderate distress | 40 (30.9 (25.7 to 36.6)) | 89 (69.1 (63.4 to 74.3)) | 129 (100.0) | |
| High distress | 54 (25.5 (15.3 to 39.2)) | 159 (74.5 (60.8 to 84.7)) | 213 (100.0) | |
| Self-management | | | | |
| Total score | | | | 0.748 |
| Good adherence | 193 (32.4 (26.2 to 39.3)) | 402 (67.6 (60.7 to 73.8)) | 595 (100.0) | |
| Poor adherence | 7 (28.3 (11.2 to 55.3)) | 18 (71.7 (44.7 to 88.8)) | 25 (100.0) | |
| Dietary control | | | | 0.386 |
| Good adherence | 174 (33.0 (26.0 to 40.8)) | 352 (67.0 (59.2 to 74.0)) | 526 (100.0) | |
| Poor adherence | 26 (27.7 (19.8 to 37.2)) | 68 (72.3 (62.8 to 80.2)) | 94 (100.0) | |
| Glucose management | | | | 0.511 |

| | | | | |
|---|---|---|---|---|
| Good adherence | 181 (32.7 (26.5 to 39.6)) | 371 (67.3 (60.4 to73.5)) | 552 (100.0) | |
| Poor adherence | 19 (28.0 (16.4 to 43.4)) | 49 (72.0 (56.6 to 83.6)) | 68 (100.0) | |
| Physician contact | | | | 0.304 |
| Good adherence | 183 (33.1 (26.3 to 40.6)) | 369 (66.9 (59.4 to 73.7)) | 552 (100.0) | |
| Poor adherence | 17 (25.1 (15.1 to 38.7)) | 51 (74.9 (61.3 to 84.9)) | 68 (100.0) | |
| Physical activity | | | | 0.019* |
| Good adherence | 110 (28.5 (22.7 to 35.2)) | 275 (71.5 (64.8 to 77.3)) | 385 (100.0) | |
| Poor adherence | 90 (38.2 (30.5 to 46.6)) | 145 (61.8 (53.4 to 69.5)) | 235 (100.0) | |
| Knowledge on diabetes | | | | 0.097 |
| Low | 62 (35.6 (27.6 to 44.6)) | 112 (64.4 (55.4 to 72.4)) | 174 (100.0) | |
| Acceptable | 68 (27.7 (20.9 to 35.7)) | 177 (72.3 (64.3 to 79.1)) | 245 (100.0) | |
| Good | 70 (34.7 (28.5 to 41.5)) | 131 (65.3 (58.5 to 71.5)) | 201 (100.0) | |

*p<0.05
